# Evidence of increasing recorded diagnosis of autism spectrum disorders in Wales, UK – an e-cohort study

**DOI:** 10.1101/2021.02.17.21250756

**Authors:** Jack F G Underwood, Marcos DelPozo-Banos, Aura Frizzati, Ann John, Jeremy Hall

## Abstract

Estimates place the prevalence of autism spectrum disorders (autism) at around 1% in the population. New services for adult diagnosis have been set up in Wales, UK, at a time of rising awareness of the spectrum of autism experiences, however no studies have examined adult autism prevalence in Wales. In this study we use an anonymised e-cohort comprised of healthcare record data to produce all-age estimates of prevalence and incidence of recorded autism for the years 2001-2016. We found the overall prevalence rate of autism in healthcare records was 0.51%. The number of new-recorded cases of autism increased from 0.188 per 1000 person-years in 2001 to 0.644 per 1000 person-years in 2016. The estimate of 0.51% prevalence in the population is lower than suggested by population survey and cohort studies study methodologies, but comparable to other administrative record study estimates. Rates of new incident diagnoses of autism saw a >150% increase in the years 2008-2016, with a trend towards more diagnoses in those over 35 and an eightfold increase in diagnoses in women from 2000-2016. Our study suggests that while the number of people being diagnosed with autism is increasing, many are still unrecognised by healthcare services.

**Lay Abstract:** Autism spectrum disorders (autism) are thought to be relatively common, with analyses estimating 1% in the population could meet diagnostic criteria. New services for adult diagnosis have been set up in Wales, UK, however no studies have examined for the proportion of adults with autism in Wales. In this study we take anonymised healthcare record data from more than 3.6 million people to produce a national estimate of recorded autism diagnoses. We found the overall prevalence rate of autism in healthcare records was 0.51%. The number of new-recorded cases of autism increased from 0.188 per 1000 person-years in 2001 to 0.644 per 1000 person-years in 2016. The estimate of 0.51% prevalence in the population is lower than suggested by population survey and cohort studies, but comparable to other administrative records. From 2001-2016 the number of autism services for adults has increased, and autism is more widely known in society, while concurrently in healthcare records there was a >150% increase autism diagnoses in the years 2008-2016. An increasing number of diagnoses were amongst women and those aged over 35. Our study suggests that while the number of people being diagnosed with autism is increasing, many are still unrecognised by healthcare services.

## Introduction

Estimates of the prevalence of autism spectrum disorders (hereafter autism) suggest rates of around 1% in the general population, but with much debate and the suggestion that not all individuals are identified (Brugha et al., 2011, 2016; Chiarotti & Venerosi, 2020; Fombonne et al., 2021; Lyall et al., 2017). The majority of studies of autism prevalence have been conducted in birth or child cohorts using a variety of epidemiological methodologies (Elsabbagh et al., 2012; Fombonne et al., 2021). These have provided varying estimates with some suggesting increasing prevalence and incidence of autism (Idring et al., 2015; Maenner et al., 2020). This potentially reflects greater societal recognition and awareness of autism beyond historic descriptions associated with intellectual disability to the full spectrum of experience (Elsabbagh et al., 2012; Russell et al., 2015). A recent systematic review and meta-analysis confirmed that in the highest quality studies a reliable estimate of male to female ratio was around 3:1, although there is increased recognition that women and girls with autism are underserved and unrecognised based on current diagnostic criteria and service provision (Lockwood Estrin et al., 2020; Loomes et al., 2017). Epidemiological studies of prevalence of autism specifically in adults are scarce, limited to pioneering work by Brugha using the UK Adult Psychiatric Morbidity Survey population case-register, a 2019 study using administrative data of autism prevalence in the United States Medicaid Program, and a 2021 study using the UK General Practice Research Database (Brugha et al., 2016; Jariwala-Parikh et al., 2019; Russell et al., 2021).

In Wales 2016 saw the launch of a new national Integrated Autism Service (IAS) as part of the Autistic Spectrum Disorder Strategic Action Plan from Welsh Government, an example of the development of services to improve provision of autism diagnosis to adults who are more likely to be undiagnosed or experience misdiagnosis (Au-Yeung et al., 2019; Holtom et al., 2019; Lai & Baron-Cohen, 2015). The global burden of autism is unknown, but in the United States and United Kingdom the annual cost is estimated in billions, and earlier identification and assistance enables adaptation and support whilst improving social communication and reducing anxiety (Buescher et al., 2014; Jariwala-Parikh et al., 2019; Lai et al., 2014). No prior whole population analyses of autism rates have been performed in Wales, with only estimates based on the Pupil Level School Age Census of Special Educational Needs and a 1988-2004 clinical registry study of children from the Rhondda and Taff Ely areas of South Wales evident in the literature (Holtom et al., 2019; Latif & Williams, 2007). Establishing incidence data on a population level offers the opportunity to inform future healthcare policy and provision, leading to improvements in quality of life for autistic people across the lifespan. To address this need we used anonymised healthcare record data from the SAIL Databank to perform an e-cohort study establishing the incidence and prevalence rates of autism diagnosed in Wales from 2001-2016.

## Methodology

### Data sources, Population and Settings

This was a retrospective population based electronic cohort study. Data was sourced from the Secure Anonymised Information Linkage (SAIL) Databank (www.saildatabank.com) (Lyons et al., 2009). SAIL is a data repository of anonymised person based linkable data from healthcare and public settings. Healthcare in Wales is provided through defined geographical catchment areas, known as Local Health Boards, under the National Health Service (NHS), whilst social care is planned and commissioned by 22 local government authorities defined by geographic area. All health boards utilise an integrated national electronic record system, the Welsh Clinical Portal, whilst individual general practices (primary care) operate their own electronic record systems. A central ‘spine’ provides personal identifier information to clinical and social care services to integrate identification. Autism diagnostic services prior to 2016 were independently commission by health boards, usually through paediatric or child and adolescent mental health care services for those under 18 years of age, and through community mental health teams for those 18 or older. Clinical encounters in secondary care are routinely recorded within the Welsh Clinical Portal, with letters then sent to individuals’ general practitioners and integrated into individuals notes held on file. Participating services submit their primary care (General Practice) clinic electronic health records, electronic hospital administration and patient health records, and central health and social care electronic administration records to SAIL, where it is anonymised at inclusion (*SAIL Databank - The Secure Anonymised Information Linkage Databank*, n.d.). The policies, permissions, processes, structures and controls applied to manage the SAIL databank are described elsewhere (Ford et al., 2009; Jones et al., 2019; Lyons et al., 2009). Data access was approved by the SAIL independent Information Governance Review Panel in 2018 (application 0843). No community members were involved in this study.

This study used the Welsh Longitudinal General Practice dataset (WLGP) and PEDW (Patient Episode Database for Wales) for extracting relevant diagnostic data, and the Welsh Demographic Service Dataset (WDSD), which includes anonymised demographics, geographic index (LSOA = Lower Layer Super Output Area index 2011) and deprivation data (Welsh Index of Multiple Deprivation quintile 2011). The WDSD is an administrative register of all individuals in Wales to use NHS services, including anonymised demographics and general practice registration. The PEDW dataset comprises hospital admissions (inpatient and day case) clinical information for all NHS Wales hospitals. At the time of the analysis 333 (of 432, 77%) general practices in Wales were supplying their data to SAIL for the WLGP dataset, covering 79% of the population. Anonymised information held in the SAIL Databank includes information pertaining to individuals who have moved into or away from the recruitment areas, or who have been born or died over the study period, reflecting population turnover. In total records for 3.63 million individuals from the Welsh population over the period 2000-2016 were examined for inclusion in the study.

Individuals with autism were identified in the WLGP and PEDW datasets using ICD (International Classification of Diseases) version 10 codes and Read Codes v2. This methodology has been validated in previous studies of other psychiatric disorders (Lloyd et al., 2015). The cohort utilised the ICD-10 codes under F84. These codes have been used in previous comparable health record linkage study designs of autism selecting for the maximum sensitivity of autistic phenotypes (Brooks et al., 2021; Idring et al., 2015; Jariwala-Parikh et al., 2019; Skonieczna-Żydecka et al., 2017). The following Read Codes v2 were selected to mirror the ICD-10 diagnostic coding: 1J9.., E140., E1400, E1401, E140z, Eu840, Eu841, Eu845, E141., Eu84., Eu842, Eu843, Eu844, Eu845, Eu84y, and Eu84z (Appendix A). Data in the SAIL databank are held in a central repository, requiring extraction and interrogation using structured query language (SQL DB2). SQL code was written to identify eligible individuals from the databank based on presence of the inclusion criteria codes in the WLGP and PEDW datasets. WDSD data were used to prevent multiple counting of cases through location change and for recording of deaths. No deprivation, race/ethnicity or geographic data were extracted for this study. Specific data on socioeconomic status and educational attainment were not available. There were no age or intellectual functioning exclusion criteria for this study.

### Statistical Analysis

Analysis of extracted outputs were processed utilising IBM SPSS Statistics 23 for Window (IBM Corp. Released, 2015) and R version 4.0.3 (R Core Team, 2020) utilising the “Tidyverse” packages (Wickham et al., 2019). For analysis of recorded incidence and prevalence, the denominator was calculated annually utilising all individuals alive and living in Wales and registered with a GP providing data to SAIL between 1st of January 2000 and 31st of December 2016. Prevalence of recorded autism and annual rates of first ever incident recorded autism were calculated for the whole 16-year period or until death, whichever was earlier. Prevalence was defined as any entry of autism in SAIL records ever, including diagnosis prior to 2000. A new incident episode was defined as an entry in the records with no previous entry of that problem ever recorded and with at least six months’ worth of WLGP data prior to that diagnosis. Annual first ever incident rates were calculated per person-years at risk, where the proportion of data held for each year for each individual was used to give an accurate estimate of record coverage. Person-time at risk was calculated using the start of each year (1^st^ January) or start of registration (plus six months), whichever later. End date was earliest of: date of leaving practice supplying SAIL, date of death, or end of year (31^st^ December).

## Results

### Autism Incidence and Prevalence

The prevalence of autism recorded in electronic health records increased across the study period of 2001-2016, from 0.189 (95% CI 0.170-0.210) per 1000 people in 2001 to 5.068 (95% CI 4.979-5.159) per 1000 people in 2016 (Table 1), reflecting cumulative incidence across the dataset. Prevalence of autism diagnosis in male electronic healthcare records prevalence rose from 0.324 (95% CI 0.289-0.364) per 1000 people in 2001 to 7.995 (95% CI 7.836-8.156) per 1000 people in 2016. Prevalence of autism diagnosis in female healthcare records rose from 0.056 (95% CI 0.043-0.074) per 1000 people in 2001 to 2.149 (95% CI 2.069-2.234) per 1000 people in 2016.

**Table 1:** Annual prevalence of record of autism diagnosis on health care datasets in the SAIL Databank over period 2001-2016

| *Year* | *Number of male cases* | *Number of males in SAIL* | *Prevalence rate per 1000* | *95% CI* | *Number of female cases* | *Number of females in SAIL* | *Prevalence rate per 1000* | *95% CI* | *Total number of cases* | *Total individuals in SAIL* | *Prevalence rate per 1000* | *95% CI* |
| --- | --- | --- | --- | --- | --- | --- | --- | --- | --- | --- | --- | --- |
| 2001 | 286 | 881,860 | 0.324 | 0.289-0.364 | 51 | 903,277 | 0.056 | 0.043-0.074 | 337 | 1,785,137 | 0.189 | 0.170-0.210 |
| 2002 | 591 | 950,872 | 0.622 | 0.573-0.674 | 122 | 973,633 | 0.125 | 0.105-0.150 | 713 | 1,924,505 | 0.370 | 0.344-0.399 |
| 2003 | 1,093 | 1,049,529 | 1.041 | 0.982-1.105 | 239 | 1,070,509 | 0.223 | 0.197-0.253 | 1,332 | 2,120,038 | 0.628 | 0.595-0.663 |
| 2004 | 1,505 | 1,081,559 | 1.392 | 1.323-1.464 | 319 | 1,101,720 | 0.290 | 0.259-0.323 | 1,824 | 2,183,279 | 0.835 | 0.798-0.875 |
| 2005 | 1,950 | 1,107,375 | 1.761 | 1.685-1.841 | 419 | 1,126,163 | 0.372 | 0.338-0.409 | 2,369 | 2,233,538 | 1.061 | 1.019-1.104 |
| 2006 | 2,357 | 1,122,794 | 2.099 | 2.016-2.186 | 499 | 1,138,141 | 0.438 | 0.402-0.479 | 2,856 | 2,260,935 | 1.263 | 1.218-1.310 |
| 2007 | 2,835 | 1,133,420 | 2.501 | 2.411-2.595 | 573 | 1,146,336 | 0.500 | 0.461-0.542 | 3,408 | 2,279,756 | 1.495 | 1.446-1.546 |
| 2008 | 3,275 | 1,143,999 | 2.863 | 2.767-2.962 | 676 | 1,153,490 | 0.586 | 0.544-0.632 | 3,951 | 2,297,489 | 1.720 | 1.667-1.774 |
| 2009 | 3,783 | 1,147,285 | 3.297 | 3.194-3.404 | 785 | 1,154,381 | 0.680 | 0.634-0.729 | 4,568 | 2,301,666 | 1.985 | 1.928-2.043 |
| 2010 | 4,321 | 1,148,708 | 3.762 | 3.651-3.875 | 918 | 1,154,189 | 0.795 | 0.746-0.818 | 5,239 | 2,302,897 | 2.275 | 2.214-2.337 |
| 2011 | 4,934 | 1,154,644 | 4.273 | 4.156-4.394 | 1,052 | 1,159,066 | 0.908 | 0.854-0.964 | 5,986 | 2,313,710 | 2.587 | 2.523-2.653 |
| 2012 | 5,603 | 1,155,864 | 4.847 | 4.722-4.976 | 1,239 | 1,158,980 | 1.069 | 1.011-1.130 | 6,842 | 2,314,844 | 2.956 | 2.887-3.026 |
| 2013 | 6,385 | 1,157,963 | 5.514 | 5.381-5.651 | 1,458 | 1,163,568 | 1.253 | 1.190-1.319 | 7,843 | 2,321,531 | 3.378 | 3.305-3.454 |
| 2014 | 7,336 | 1,172,841 | 6.255 | 6.114-6.399 | 1,782 | 1,177,854 | 1.513 | 1.444-1.585 | 9,118 | 2,350,695 | 3.879 | 3.800-3.959 |
| 2015 | 8,269 | 1,176,629 | 7.051 | 6.878-7.180 | 2,119 | 1,180,120 | 1.796 | 1.720-1.874 | 10,415 | 2,356,749 | 4.419 | 4.335-4.504 |
| 2016 | 9,523 | 1,191,143 | 7.995 | 7.836-8.156 | 2,566 | 1,194,013 | 2.149 | 2.068-2.234 | 12,089 | 2,385,156 | 5.068 | 4.979-5.159 |

*SAIL = Secure Anonymised Information Linkage Databank.*

The yearly incidence for new recording of autism diagnosis in electronic health records from 0.188 (95% CI 0.169-0.209) per 1000 person-years in 2001, up to a maximum of 0.644 (95% CI 0.612-0.677) per 1000 person-years in 2016 (Table 2), a 3.4-fold increase. This change comprised an increase in recorded incidence in both male and female individuals, from 0.322 (95% CI 0.286-0.361) and 0.057 (95% CI 0.043-0.075) per 1000 person-years at risk up to 0.945 (95% CI 0.891-1.003) and 0.345 (95% CI 0.313-0.380) per 1000 person-years respectively for 2016.

**Table 2:** Annual incidence of record of autism diagnosis on health care datasets in the SAIL Databank over period 2001-2016

| *Year* | *Incident male cases* | *Male person-years in SAIL* | *Male incidence rate per 1000 person-years* | *95% CI* | *Incident female cases* | *Female person-years in SAIL* | *Female incidence rate per 1000 person-years* | *95% CI* | *Total incident cases* | *Total person-years in SAIL* | *Cumulative incidence rate per 1000 person-years* | *95% CI* |
| --- | --- | --- | --- | --- | --- | --- | --- | --- | --- | --- | --- | --- |
| 2001 | 282 | 876,919.13 | 0.322 | 0.286-0.361 | 51 | 897,691.11 | 0.057 | 0.043-0.075 | 333 | 1,774,610.24 | 0.188 | 0.169-0.209 |
| 2002 | 271 | 945,126.81 | 0.287 | 0.255-0.323 | 63 | 945,126.81 | 0.065 | 0.052-0.085 | 334 | 1,912,550.35 | 0.175 | 0.157-0.194 |
| 2003 | 403 | 1,042,719.27 | 0.386 | 0.351-0.326 | 99 | 1,063,617.87 | 0.093 | 0.076-0.113 | 502 | 2,106,337.14 | 0.238 | 0.218-0.260 |
| 2004 | 382 | 1,074,281.41 | 0.356 | 0.322-0.393 | 74 | 1,094,790.53 | 0.068 | 0.054-0.085 | 456 | 2,169,071.94 | 0.210 | 0.192-0.230 |
| 2005 | 410 | 1,099,552.14 | 0.373 | 0.338-0.411 | 94 | 1,118,843.52 | 0.084 | 0.069-0.103 | 504 | 2,218,395.66 | 0.227 | 0.208-0.348 |
| 2006 | 394 | 1,114,591.24 | 0.353 | 0.320-0.390 | 78 | 1,130,926.23 | 0.069 | 0.055-0.086 | 472 | 2,245,507.47 | 0.210 | 0.192-0.230 |
| 2007 | 454 | 1,124,613.57 | 0.404 | 0.368-0.443 | 72 | 1,138,830.32 | 0.063 | 0.050-0.080 | 526 | 2,263,443.89 | 0.232 | 0.213-0.253 |
| 2008 | 425 | 1,134,605.58 | 0.375 | 0.341-0.412 | 106 | 1,145,951.41 | 0.092 | 0.077-0.112 | 531 | 2,280,556.99 | 0.233 | 0.214-0.253 |
| 2009 | 488 | 1,137,293.25 | 0.429 | 0.393-0.469 | 105 | 1,146,686.39 | 0.092 | 0.076-0.111 | 593 | 2,283,979.64 | 0.260 | 0.240-0.281 |
| 2010 | 546 | 1,138,448.90 | 0.480 | 0.441-0.522 | 134 | 1,146,347.58 | 0.117 | 0.099-0.138 | 680 | 2,284,796.48 | 0.298 | 0.276-0.321 |
| 2011 | 593 | 1,143,509.86 | 0.519 | 0.478-0.562 | 130 | 1,151,224.22 | 0.113 | 0.095-0.134 | 723 | 2,294,734.08 | 0.315 | 0.293-0.339 |
| 2012 | 646 | 1,143,863.93 | 0.565 | 0.523-0.610 | 177 | 1,150,621.65 | 0.154 | 0.133-0.178 | 823 | 2,294,485.58 | 0.359 | 0.334-0.384 |
| 2013 | 755 | 1,144,867.07 | 0.659 | 0.614-0.708 | 219 | 1,154,565.52 | 0.190 | 0.166-0.217 | 974 | 2,299,432.59 | 0.424 | 0.398-0.451 |
| 2014 | 846 | 1,158,887.39 | 0.730 | 0.682-0.781 | 284 | 1,168,928.58 | 0.243 | 0.216-0.273 | 1,130 | 2,327,815.96 | 0.485 | 0.458-0.515 |
| 2015 | 942 | 1,161,144.89 | 0.811 | 0.761-0.865 | 308 | 1,169,864.99 | 0.263 | 0.235-0.294 | 1,250 | 2,331,009.88 | 0.536 | 0.507-0.567 |
| 2016 | 1,110 | 1,174,191.93 | 0.945 | 0.891-1.003 | 408 | 1,183,570.16 | 0.345 | 0.313-0.380 | 1,518 | 2,357,792.09 | 0.644 | 0.612-0.677 |

*SAIL = Secure Anonymised Information Linkage Databank.*

The number of new incident recorded diagnosis of autism on males’ healthcare records in SAIL increased by 3.94 times over the study period, from 282 cases on 2001 records, to 1,110 cases on 2016 records (Table 2). This increase was half the eightfold rate increase seen amongst female healthcare records, from 51 new diagnoses recorded in 2001 to 408 in 2016. This coincided with an increase in the number of diagnoses made in adulthood. In 2001, 19 (6.74%) males over the age of 35 had a new diagnosis of autism recorded on their healthcare record, by 2016 this had increased to 100 (9.01%) males over the age of 35 with a new diagnosis of autism recorded in that year (Figure 1). This trend was seen across both sexes.

**Figure 1:**
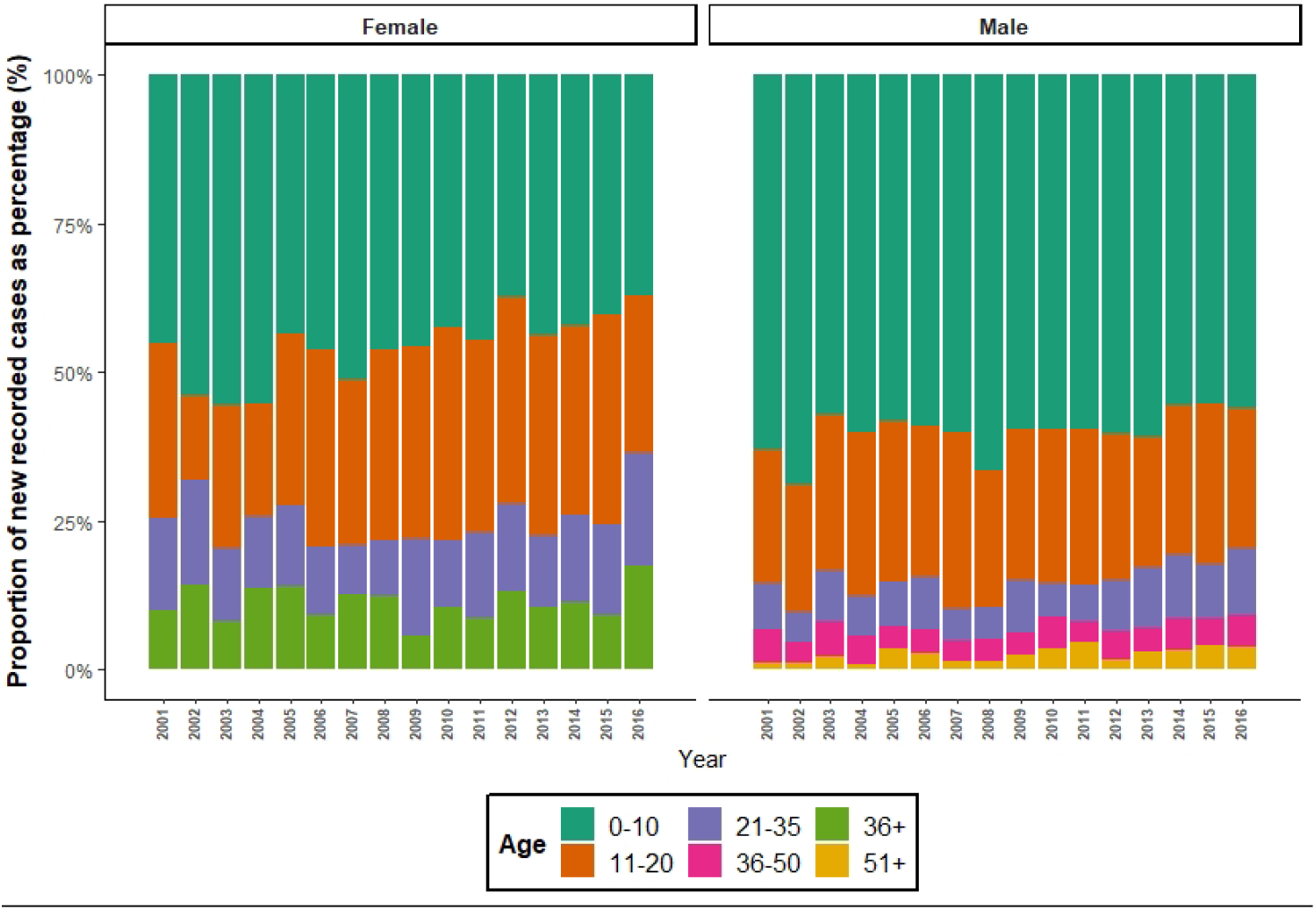
Proportion of incident recorded cases divided by sex and age-grouping over assessed years (2001-2016) Number of new recorded cases of autism recorded on healthcare record plotted by sex and age grouping plotted over assessed period (2001-2016) in SAIL Databank. Age groupings to meet minimum sample size reporting guidelines for SAIL databank.

## Discussion

In this large population-based electronic-cohort study we draw on anonymised healthcare record data from General Practice and hospital encounters for over 3.6 million individuals to produce the first national all-age estimates of recorded autism prevalence and incidence for Wales. We found a recorded prevalence of autism of 5.068 (95% CI 4.979-5.159) per 1000 people in 2016 equivalent to 0.51% of the population, and a 3.4-fold increase in recorded autism incidence over the study period from 0.188 (95% CI 0.169-0.209) per 1000 person-years in 2001, up to a maximum of 0.644 (95% CI 0.612-0.677) per 1000 person-years in 2016.

We demonstrate an increase in recorded incidence of autism diagnosis, particularly in the eight-year period from 2008-2016. During this time new recorded diagnoses increased by >150%. Despite this, the absolute recorded incidence rate remains low. Multiple groups have reported that rates of autism diagnosis are increasing over time (Lai et al., 2014; Russell et al., 2015). A previous study of comparative data from the General Practice Research Database (GPRD) demonstrated that the incidence rate amongst children aged 8 years during an overlapping time period (2001-2010) remained static, however a more recent analysis across the whole lifespan and for a wider time period (1998-2018) in the same dataset by Russell *et al* found that there was a 787% exponential increase in recorded incidence of autism diagnoses (Russell et al., 2021; Taylor et al., 2013). The authors of the 2021 study found the increase in diagnoses was greater in females than in males and concluded that this was likely due to greater awareness and availability of diagnosis (Russell et al., 2021). We would therefore suspect that the increase in incident recorded diagnoses in the present study represents better identification and provision for diagnosis of autism in the healthcare system. This particularly noticeable amongst the older population and women, where there has been considerable systematic diagnostic bias, again replicating Russell *et al*’s findings (Loomes et al., 2017; Russell et al., 2021). We observed a trend towards more diagnoses being made amongst adults over 35, and eight times the number of women diagnosed with autism in 2016 than were in 2001. We, like others, would suggest that this increase is due to recognition, increased societal awareness, changes in practice and improved availability of diagnostic services particularly for adults with autism (Brugha et al., 2016; Elsabbagh et al., 2012; Russell et al., 2015).

Estimates point to a prevalence of autism in the general population around 1% (Abdallah et al., 2011; Baio et al., 2018; Chiarotti & Venerosi, 2020; Fombonne et al., 2021; Lord et al., 2020; Lyall et al., 2017; Russell et al., 2015; Taylor et al., 2013). Prior studies of autism prevalence have examined a number of birth or school cohorts, CDC surveillance data on children’s health records in the US, and the General Practice Research Database (GPRD) for general practice health records of children in the UK (Baio et al., 2018; Elsabbagh et al., 2012; Fuentes et al., 2020; Russell et al., 2015; Taylor et al., 2013; Thomaidis et al., 2020). Brugha *et al* were reportedly the first to ascertain data for adults, finding a prevalence of 0.98-1.1% using a national stratified survey methodology (Brugha et al., 2011, 2016). Further studies have corroborated the ∼1% estimate, with Fombonne *et al*’s 2021 review paper citing a median prevalence in 26 high income countries of 0.97% (Chiarotti & Venerosi, 2020; Fombonne et al., 2021). Our prevalence estimate of 5.068 (95% CI 4.979-5.159) per 1000 people is lower than these epidemiological studies. This may be due to methodological differences reflecting the under-identification and under-reporting of autism in healthcare records, or due to differences of sample population; most published autism prevalence studies are in children, with few studies of autism prevalence in adults or across the lifespan as in this study.

Our prevalence figure is more comparable to other healthcare record and administrative data analyses. Of these Jariwala-Parikh *et al’s* 2019 study of autism prevalence in the US Medicaid programme is one of the only few examining adults, finding an annual prevalence of autism diagnosis of 3.66 per 1000 in 2008 (Jariwala-Parikh et al., 2019). Studies in Europe using administrative data and with health care systems similar to Wales have produced estimates of between 3.5-5.8/1000 children in Poland, 3.8/1000 in Germany, 3.6/1000 in France, 5.8-15/1000 in Greece, 4.0-24.6/1000 in Sweden and 5.9/1000 in Spain (Bachmann et al., 2016; Delobel-Ayoub et al., 2015; Fuentes et al., 2020; Idring et al., 2015; SkoniecznaZydecka et al., 2017; Thomaidis et al., 2020). The majority of these studies were conducted in child cohorts, and therefore it would be expected prevalence would increase with annual incidence of new diagnosis into adulthood. In our cohort variance due to population turn-over means that while annual recorded autism prevalence increases, this is not equal to the growth of annual incidence. Annual recorded incidence likely exceeds any change in autism population prevalence, therefore demonstrating under-identification and reporting.

This study utilised the SAIL Database, a nationally-representative, high quality anonymised e-cohort of >3.6 million individuals (Lyons et al., 2009). We reviewed General Practice and Hospital Encounter datasets, comprising ICD-10 and Read v2 Codes for diagnoses under analysis. As an e-cohort study, exposure and outcome classification errors cannot be excluded as we are unable to verify the diagnoses. Electronic records of General Practice data prior to 2000 are sparsely held in the SAIL Databank, and therefore prevalent cases prior to 2000 are likely missing from this estimate. This explains the dramatic increase from very low baseline of our reported prevalence. It is likely our study misses individuals with autism in the community who have not come into contact with healthcare services resulting in their diagnosis not being on record. Prior to 2016 autism diagnostic services in Wales were organised by individual health board, and disparate in availability and structure (Holtom et al., 2019). Anecdotal reports suggest many adults sought private diagnoses due to lack of provision, which may not have reported findings into NHS healthcare services. Our study is therefore likely to be an underestimation. Diagnostic overshadowing, the practice of one diagnosis taking primacy and leading to underdiagnosis of co-occurring conditions, is also recognised to be a problem within autism (Hollocks et al., 2018). This study used the ICD-10 code F84 to define cases of autism, along with comparable identifiers from Read Codes v2. This methodology has been used by comparable studies to maximise sensitivity to the autism phenotype, however it does include diagnoses such as Rett syndrome which are syndromes associated with specific causal factors which may not be representative of autism as a whole (Idring et al., 2015; Jariwala-Parikh et al., 2019; Skonieczna-Żydecka et al., 2017). The definition of autism case status remains a challenge among epidemiological studies, with no uniform method among published studies (Fombonne et al., 2021). These limitations may compound to generate errors in our estimates and contribute to lower figures, although our large sample size and replication of comparable results would suggest these errors are minimal. We have attempted to compensate through use of both primary and secondary care data, and these limitations are common to e-cohort and record linkage studies which depend upon individuals’ attendance, recognition and recording to be accurate.

In conclusion, in our national population sample we found an increase in annual recorded incidence over the study period to 2016. This included increases in diagnosis of females and those over age 35. This is consistent with improved societal recognition and access to diagnostic services resulting in greater rates of diagnosis; incident cases not picked up in childhood being diagnosed as adults. Despite this we found lower prevalence rates than demonstrated in previous epidemiological survey and birth cohort studies, comparable to other health record and administrative study methodologies, suggesting that a large proportion of those adults with autism are not recognised by the healthcare system. Increased rates of diagnosis and more prevalent autism in the community necessitates increased funding for specialist services to enable autistic adults to receive any support they require.

## Data Availability

Data referred to is held within the SAIL Databank and governed by data access approval systems.

## Funding

This project was funded by a Wellcome Trust ISSF Clinical Primer award, and subsequent Wellcome Trust GW4-CAT Clinical Doctoral Fellowship (222849/Z/21/Z) to JFGU. A CC BY or equivalent licence is applied to the Authors Accepted Manuscript arising from this submission, in accordance with the grant’s open access conditions. JH, AJ, MDPB and JFGU are supported by Healthcare Research Wales through the National Centre for Mental Health (NCMH) (CA04). JH and JFGU are supported by an MRC Pathfinder Grant (MC_PC_17212), and MDPB, AF and AJ are supported by an MRC Pathfinder Grant (MC_PC_17211).

## Contributors

JFGU principally designed the study, acted as principal investigator, statistically analysed the data and wrote the first draft of the manuscript with input from AF. AF, MDPB, AJ and JH all contributed to the design of the study. AF and MDPB co-assisted with data retrieval and analysis. All authors critically read the manuscript and contributed to the data interpretation and writing. All authors approved the final version of the manuscript. The study protocol and summary data may be requested from JFGU. The study protocol and summary data may be requested from JFGU.

## Declaration of interests

JH has received grants from Takeda for research work unrelated to this project. The other authors declare no competing interests.

## Notes

### Author Declarations

Data access was approved by the SAIL independent Information Governance Review Panel in 2018 (application 0843). The policies, permissions, processes, structures and controls applied to manage the SAIL databank are described elsewhere (John et al., 2018).

### Summary of Updates

Extensive revision up to extent of Authors Accepted Manuscript for submission to Autism.

