## Appendix A for "Evidence of increasing recorded diagnosis of autism spectrum disorders in Wales, UK – an e-cohort study"

**Coding utilised for identification of autism spectrum disorder phenotype in SAIL**

**International Classification of Diseases – 10^th^ revision:**

- F84 – Pervasive developmental disorders, containing:
  - F84.0 – Childhood autism
  - F84.1 – Atypical autism
  - F84.2 – Rett’s syndrome
  - F84.3 – Other childhood disintegrative disorder
  - F84.4 – Overactive disorder associated with mental retardation and stereotyped movements
  - F84.5 – Asperger’s syndrome
  - F84.8 – Other pervasive developmental disorders
  - F84.9 – Pervasive developmental disorders, unspecified

**Read Codes v2:**

The following Read Codes v2 were selected to mirror the ICD-10 diagnostic coding:

- E140. – Autism/ infantile autism
- E1400 – Active infantile autism
- E1401 – Residual infantile autism
- E140z – Infantile autism NOS
- Eu840 – Childhood autism/ infantile autism
- Eu841 – Atypical autism
- Eu845 – Asperger syndrome
- 1J9.. – Suspected autism
- E141. – Childhood disintegrative disorder
- Eu84. – Pervasive developmental disorder
- Eu842 – Rett syndrome
- Eu843 – Other childhood disintegrative disorder
- Eu844 – Overactive disorder associated with mental retardation and stereotyped movements
- Eu845 – Asperger syndrome
- Eu84y – Other pervasive developmental disorder
- Eu84z – Pervasive developmental disorder, unspecified
